# Validation of the Fremantle Perineal Awareness Questionnaire (FrePAQ) in women with Chronic Pelvic Pain

**DOI:** 10.64898/2026.06.05.26354913

**Authors:** Jilly Bond, Neil E O’Connell, Benedict M Wand, K. Jane Chalmers, Elmar Kal

## Abstract

**Background:** Chronic pelvic pain (CPP) affects up to 26% of women worldwide. While its pathophysiology is poorly understood, disturbances in body perception have been identified in various similar chronic musculoskeletal disorders. The Fremantle Perineal Awareness Questionnaire (FrePAQ) is a novel tool designed to specifically assess disturbed body perception in the pelvic region, but its structural validity and reliability require formal evaluation.

**Methods:** Patient partners with lived experience contributed to study design. Participants with (n=417 and without (n=277) chronic pelvic pain completed the FrePAQ at baseline, as well as one week later. We assessed the validity and reliability of the FrePAQ following COSMIN guidelines for Classical Test Theory.

**Results:** The validated FrePAQ comprises a two-factor model, with a six-item Distress & Disconnection (D&D) subscale and a two-item Size & Shape (S&S) subscale. Confirmatory analysis showed excellent fit (CFI = .988; RMSEA = .048) and measurement invariance between diagnostic groups. Internal consistency was high (α = .838 CPP; .819 controls). Test–retest reliability was high for D&D (ICC = .863) and acceptable for S&S (ICC = .695). FrePAQ scores showed a weak-moderate correlation with pain scores (r = .234–.255), psychological distress (r = .226–.443), and functional impact (r = .172–.295), particularly for the D&D subscale.

**Conclusion:** The FrePAQ is a reliable and valid instrument to measure perineal perceptual disturbances in CPP. Future research will evaluate the tool’s potential to support phenotyping and guide individualised interventions. Improved understanding of body perception disturbance in CPP can enhance diagnosis and treatment precision.

**Perspective:** This article validates the Fremantle Perineal Awareness Questionnaire (FrePAQ), the first measure of perineal body perception disturbance in chronic pelvic pain. The findings support body perception disturbance as a clinically relevant feature of chronic pelvic pain and provide a foundation for future research exploring its role in phenotyping, treatment selection, and treatment response.

## Introduction

Chronic pelvic pain (CPP) affects up to 26% of women worldwide and contributes an estimated $2.8 billion in annual healthcare costs^1^. It is a leading health issue among reproductive-aged women^2^, and is defined as ongoing or recurrent pelvic pain lasting at least three months, or six months if cyclical^3^. The condition is complex, involving dysfunctions related to the pelvic floor, urinary, bowel, or gynaecological systems and commonly presents with comorbidities such as fibromyalgia, migraine, sleep disorders, anxiety, and depression^4^. While international guidelines recommend multimodal and multidisciplinary treatment, particularly physiotherapy^3,5,6^, a limited understanding of the causes and development of CPP subtypes continues to hinder the creation of more targeted care^7^. It is therefore crucial to improve our understanding of CPP to enhance diagnostic accuracy and treatment precision.

Body perception disturbance (BPD) refers to an altered perception and experience of one’s own body, characterised by distortions in the perceived size, shape, position, and ownership of a body part, alongside a disrupted sense of agency over that region^8^. BPD is suggested to play a role in the maintenance and expression of other chronic pain conditions through disruption in sensorimotor and cognitive-affective integration^9–13^, and may also be an important mechanism in the manifestation of CPP. For instance, qualitative studies have highlighted that women with CPP describe disconnection from, altered awareness of, and a lack of perceived control over the pelvic region^14–19^. Further, a recent scoping review identified emerging, but inconsistent, evidence of BPD across 11 CPP conditions ^20^.

Interventions that aim to target BPD in other chronic pain conditions (e.g., graded sensorimotor retraining and graded motor imagery) have shown potential to reduce pain and disability^21,22^. These approaches have recently been applied to CPP^23–25^, with small improvements reported in pain and pelvic floor muscle function in both pain-free women and those with Genito-Pelvic Pain/Penetration Disorder or CPP. However, the translation of these interventions from other chronic pain conditions to CPP currently lacks a strong theoretical and empirical foundation, and it remains unclear whether the underlying mechanisms are comparable. Developing a more detailed and nuanced understanding of the presentation of BPD in CPP could help to describe clinically informative phenotypes which may then allow for more targeted and effective treatments^26^.

Studies of BPD in pelvic pain have predominantly focussed on sensory function using quantitative sensory testing of the region, while motor alterations have received comparatively less attention, and there has been little focus on the resulting changes in body perception ^20^. The recently developed Fremantle Perineal Awareness Questionnaire (FrePAQ) is a novel tool designed to address this gap, by assessing alterations in the perceptual experience of the pelvic region in people with CPP^27^. Although preliminary testing showed good content and face validity, its structural validity and reliability have not yet been established. As such, this study aimed to undertake a formal clinimetric validation of the FrePAQ.

## Methods

Ethical approval for this study was provided by Brunel University of London’s College of Health, Medicine and Life Sciences Research Ethics Committee [47793-MHR-Jul/2024-51960-2]. The protocol was pre-registered through the Open Science Framework ^28^ (https://shorturl.at/M60aL).^29^

### Patient Partner and Stakeholder Involvement

An informal online advisory group informed the development of background questions, survey burden, and the relevance of measures used for construct validity. With permission, the group included four UK women with lived experience of CPP from diverse ethnic backgrounds, three representatives from pelvic pain charities who also had lived experience of CPP and a gynaecological clinical academic with specialist expertise in CPP management, CPP clinical research, and antiracist research frameworks.

### Participants

This study pragmatically focused on cisgendered women as CPP is considerably more common in cisgendered women (26.6% ^30^) than in cisgendered men (2–16% ^31^), making this group the most clinically relevant starting point for validation work. Throughout this paper, the word “women” refers specifically to cisgender women included in this sample.

To capture a broad range of CPP conditions participants were recruited internationally via social media (Facebook, Instagram, X, LinkedIn), UK pelvic pain charities supporting diverse ethnic groups, and medical and allied health clinics across Europe, Canada, Australia, and South Africa (see Supplementary material for a full list). Recruitment posters provided a link to a Jisc online survey ^32^ with participant information, eligibility screening, and consent form.

Table 1 lists the specific eligibility criteria for the CPP and control group participants. Broadly speaking, we recruited adult cisgender women either with or without self-reported symptoms of CPP as per European Association of Urology (EAU) guidelines^33^. Participants with a history of other major health conditions (e.g., active cancer), pregnancy, or who had given birth within six months were excluded.

**Table 1.**
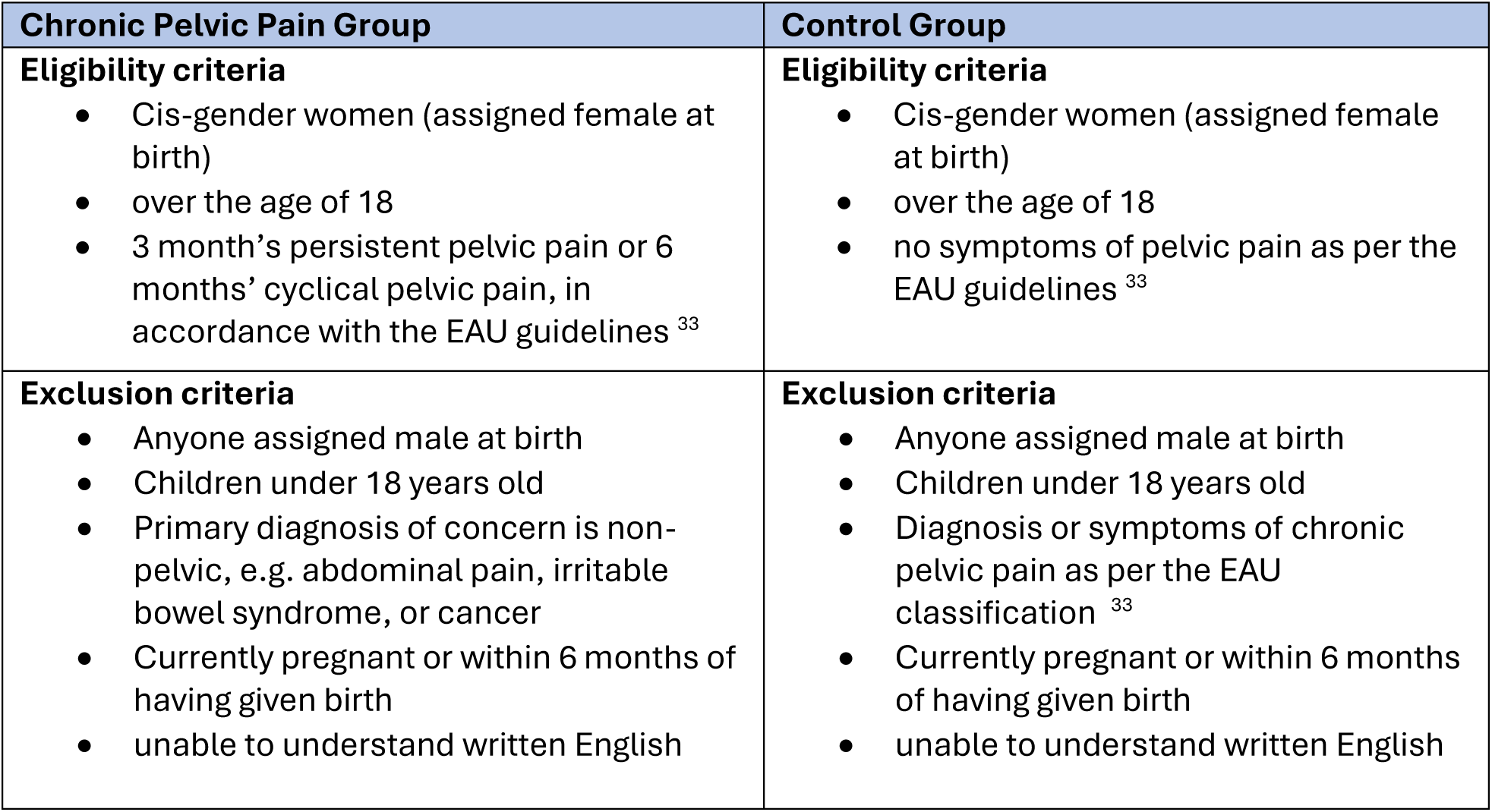
Participant inclusion and exclusion criteria.

A degree of diagnostic uncertainty was accepted as definitions of pelvic pain conditions vary internationally. We allowed participants to self-report all pelvic and non-pelvic comorbidities to better reflect the overlapping and comorbid nature of pelvic pain. We also included those without a formal diagnosis, as up to 50% of women with CPP remain undiagnosed^34^ and this group is underrepresented in the evidence base^35–37^. To address this uncertainty, participants identified their most impactful diagnosis or symptoms and indicated whether this was formally diagnosed. They were then asked to select symptoms that best matched theirs from the list below, which reflect key features of common CPP conditions based on the EAU 2023 guidelines^33^, and that we used to classify participants:

- Cyclical pain with painful cramping, pain with passing stool, pain with intercourse, bloating and fatigue (Endometriosis, Adenomyosis and Dysmenorrhoea)^38^
- Vulval pain which worsens with pressure to the area (Vulvodynia and Vestibulodynia)^39^
- Pelvic Pain associated with bladder filling and relief with bladder emptying, urge to empty the bladder and frequent bladder emptying (Bladder Pain Syndrome)^40,41^
- Pain with intercourse, vaginal block or spasm feeling with intercourse (Vaginismus, Genito-pelvic pain/penetration disorder)^42,43^

We aimed for a sample of >500 participants as this would (i) ensure that we would have 2 samples of >250 participants each for the exploratory factor analysis (EFA) and confirmatory factor analysis (CFA), and (ii) our sample would comfortably exceed the typically recommended minimal subject-to-variable ratio of 10:1^44^. For test–retest reliability analysis, n=60 “stable” participants would be required sufficient to be able to detect an ICC of 0.80 with a 95% confidence interval (CI) of 0.70–0.90. With >500 participants, we therefore anticipated to easily reach that target, assuming response rate ∼30% and >50% of those remaining ‘stable’ in the 1-week re-test interval (see below).

### Data Collection

Data were collected using Jisc Online Surveys^32^ between July and December 2024 as part of a broader study of perineal BPD in women with CPP. Participants completed the FrePAQ at Baseline (T1) and one-week later (T2; for retest reliability). A one-week interval was selected to balance the risk of recall bias associated with shorter intervals against the possibility of true clinical change over longer periods, consistent with recommendations for patient-reported outcome measures^45,46^. At T2 participants were asked whether their symptoms had “substantially worsened”, “slightly worsened”, “stayed the same”, “slightly improved” or “substantially improved”. Only participants reporting stable symptoms were included in retest reliability analyses. At T1, participants also completed questions about their demographic and clinical characteristics (e.g., age, country, immigration status, ethnicity, sexual orientation, parity, education; see supplementary material for full list of items).

### Outcome Measures

The FrePAQ was previously developed by adapting the Fremantle Back Awareness Questionnaire (FreBAQ)^47^ through an iterative three-round modified electronic Delphi involving experts and large samples of people with CPP^27^. This resulted in a 9-item questionnaire with promising face and content validity, which was used for the present study. Each item is scored from 0 (“Never”) to 3 (“Always”), with higher scores indicating more frequent symptoms (See supplementary material for the 9-item FrePAQ).

At T1, participants additionally completed self-reported outcomes for the functional impact (Pelvic Pain Impact Questionnaire; PPIQ^48^), and distress associated with CPP (Pelvic Pain Psychological Screening Questionnaire; 3PSQ^49^), alongside numerical rating scale scores of their maximum and average pain intensity (0 = no pain, 10 = as bad as it could possibly be). Note that these outcomes were completed by participants reporting CPP only.

### Data Analysis

Data were analysed using SPSS (29.0)^50^ and AMOS (29.0)^51^. Characteristics of the CPP and control groups were summarised descriptively and compared using appropriate statistical tests based on data distribution and variable type.

Validation involved the following steps. First, the behaviour of individual FrePAQ items was evaluated, including the number of missing responses (at T1), the distribution of scores across the available response options (at T1), each item’s test–retest reliability (two-way, random effect, consistency, single measures, intraclass correlation coefficient, based on data for T1 and T2). At this point we considered removing items with a relatively high number of missing responses (>5%), with a high percentage of maximal or minimal scores (>50%) or with low reliability (ICC<0.5)^52^.

Next, the T1 data were randomly split into equal Exploratory and Confirmatory subsets for analysis. We performed an exploratory factor analysis (EFA) on the Exploratory subset of T1 data using principal axis factoring and direct oblimin factor rotation for clarity and simplicity of factor loading of the whole scale^53^. Based on similar validations of other Fremantle body awareness questionnaire^54–56^ the FrePAQ was anticipated to be unidimensional. The scree plot was examined to identify the number of latent factors. Items with factor loadings <0.3, with low item-rest (r’s < 0.3) and/or high interitem correlation (r’s > 0.7) were considered for removal at this stage. We planned to rerun the EFA if an item was removed at this stage.

Subsequently, a confirmatory factor analysis (CFA) using maximum likelihood estimation was run on the Confirmatory subset of T1 data to check if the factor structure identified through EFA had sufficient model fit. Model fit was evaluated using the χ² statistic, χ² divided by degrees of freedom (χ²/df), comparative fit index (CFI), goodness-of-fit index (GFI), root mean square error of approximation (RMSEA), and standardised root mean square residual (SRMR), with the following cut-off values indicating acceptable fit: **χ**^2^ non-significance, **χ**^2^/df <3, CFI >.95, RMSEA <.05, and SRMR <.08^57^. Items were constrained to load on the underlying factors identified in the EFA, and pairs of error terms were allowed to covary.

Next, to investigate whether the FrePAQ behaves similarly in people with and without CPP, a multi-group measurement invariance analysis was performed on the total T1 data (both Exploratory and Confirmatory subsets combined). To assess whether the scale structure was similar across the groups tested we evaluated model fit when: 1) item-factor loadings were free to differ between the selected subgroups (configural invariance), 2) when item-factor loadings were held constant between groups (metric invariance), and 3) when item-factor loadings and intercepts were held constant across all groups (scalar invariance). Configural invariance was acceptable if CFI >.90, RMSEA and SRMR are both<.08, and GFI >.90. For metric and scalar invariance, changes in model fit of ΔCFI<-0.010, ΔRMSEA<0.015, and ΔSRMR<0.030 (metric invariance) or ΔSRMR<0.010 (scalar invariance) were considered acceptable^58^.

We then evaluated internal consistency (Cronbach’s Alpha, T1 data) and re-test reliability (two-way random effect consistency, single measure ICC; T1 and T2 data) for the total scale scores (or for subscales, if identified by factor analysis)^49^. For both Cronbach’s alpha and the ICC, we considered values of > 0.70 to be satisfactory^59^. Measurement error (SEM = SD × √[1−ICC]) and minimal detectable change (MDC) were assessed at the group and individual level (MDC group = SEM × 1.96 × √2 / √n; MDC individual = SEM × 1.96 × √2).

A Pearson’s r was used to evaluate construct validity, by testing the hypotheses that there will be a positive, moderate correlation between total T1 FrePAQ score and VAS pain scores, 3PSQ scores for anxiety and depression and total PPIQ score as a measure of functional impact, accepting 0.1 as small, 0.3 as moderate, and 0.5 as large^59^.

### Results

#### Participants

A total of 711 responses were received, including 431 participants with CPP and 280 healthy controls. Seventeen responses were excluded due to incomplete data, leaving a final sample of 417 participants with CPP and 277 healthy controls. Group demographics are summarised in Table 2.

**Table 2.** Summary of participant characteristics.

|  |  | **Pelvic Pain**<br>**(n=417)** | **Control**<br>**(n=277)** | **p-value** |
| --- | --- | --- | --- | --- |
| **Age** | Mean Age (M± SD (range)) | 37.7<sup>a</sup> ± 11.9 (18- 81) | 39.55<sup>a</sup> ± 11.7 (21-86) | **.017** |
| **Location** | Australia | 39 (9.4%) | 26 (9.4%) | .273 |
|  | Canada | 45 (10.8%) | 26 (9.4%) |  |
|  | Europe | 42 (10.1%) | 23 (8.3%) |  |
|  | UK | 221 (53 %) | 126 (45.5%) |  |
|  | USA | 53 (12.7%) | 56 (20.2%) |  |
|  | Other | 17 (4 %) | 20 (7.2%) |  |
| **Migration Status** | Migrant (no., (%)) | 54 (12.9%) | 50 (18.1%) | .050 |
| **Ethnicity** | Asian | 10 (2.4%) | 11 (3.9%) | .964 |
|  | Black, Caribbean or African | 2 (0.5%) | 4 (1.4%) |  |
|  | Hispanic or Latino | 6 (1.4%) | 1 (0.4%) |  |
|  | Native or Indigenous | 2 (0.5%) | 1 (0.4%) |  |
|  | White | 378 (90.6%) | 250 (90.4%) |  |
|  | Mixed or multiple ethnic groups | 16 (3.8%) | 4 (1.4%) |  |
|  | Other | 3 (0.7%) | 6 (2.1%) |  |
| **Self-reported sexuality** | Heterosexual | 344 (82.5%) | 254 (91.8%) | **<.001** |
|  | Lesbian | 9 (2.2%) | 1 (0.4%) |  |
|  | Bisexual/Pansexual | 46 (11%) | 14 (5%) |  |
|  | Asexual/Aromantic | 6 (1.4%) | 2 (0.7%) |  |
|  | Queer/other | 7 (1.7%) | 3 (1.1%) |  |
|  | Prefer not to say | 5 (1.2%) | 3 (1.1%) |  |
| **Highest Educational Attainment** | Secondary or High School | 39 (9.2%) | 16 (5.7%) | **.010** |
|  | Higher Education or Degree | 208 (49.9%) | 118 (42.5%) |  |
|  | Postgraduate Degree | 170 (40.9%) | 143 (51.8%) |  |
| **Parity** | 0 | 254 (60.9%) | 105 (37.9%) | **<.001** |
|  | 1 | 60 (14.4%) | 48 (17.5%) |  |
|  | 2+ | 103 (24.7%) | 124 (44.6%) |  |
| **Reproductive Status** | Menstruating | 292 (70.1%) | 194 (69.9%) | **.912** |
|  | Perimenopausal | 45 (10.9%) | 31 (11.2%) |  |
|  | Postmenopausal | 56 (13.5%) | 47 (17.1%) |  |
|  | Premature Ovarian Insufficiency | 23 (5.6%) | 5 (1.8%) |  |
| **Cycle Stage<sup>b</sup>** | Menstruating | 51 (12.3%) | 28 (10%) | **.788** |
|  | Week post menstrual | 60 (14.4%) | 43 (15.7%) |  |
|  | Ovulating/middle of cycle | 75 (18.1%) | 62 (22.5%) |  |
|  | Week before period | 116 (27.8%) | 67 (24.3%) |  |
|  | Unsure | 114 (27.4%) | 76 (27.5%) |  |
| **Comorbidities** | Non-pelvic pain comorbidities | 227 (54.4%) | 88 (31.8%) | **<.001** |
| **Medications** | Non-pelvic pain medications | 2.33 (SD 2.51) | 1.72 (SD 1.62) | **.085** |
| **Physical Activity** | Regularly Exercise | 297 (71.2%) | 235 (85.2%) | **<.001** |
| **Weekly Exercise<sup>c</sup>** | 0-2 hours | 80 (26.3%) | 51 (21.5%) | **.088** |
|  | >2-4 hours | 110 (36.2%) | 89 (37.6%) |  |
|  | >4-6 hours | 67 (22%) | 50 (21.1%) |  |
|  | >6 hours | 47 (15.5%) | 47 (19.8%) |  |

NB: <sup>a</sup>1 missing case for each group; <sup>b</sup> only completed by those menstruating regularly: 118 missing cases for the CPP group, 77 missing cases for the control group; <sup>c</sup> only completed by those who identify as physically active, 113 missing cases for CPP group and 40 missing cases for control group;

More than 90% of participants in both groups self-reported as White, and, while participants reported living in 34 different countries, in both groups most were from the UK (53% CPP, 45.5% controls) and USA (12.7% CPP, 20.2% controls). The CPP and control groups did differ in some respects. For one, the CPP participants were younger (mean difference 2.2 years; p=.017), less often identified as heterosexual (82.5% in CPP, 91.8% in controls; p < .001), and reported slightly lower educational attainment (p = .010). The CPP group also had significantly lower parity, with 60.9% nulliparous versus 37.9% in the control (p<.001), but overall, there was no statistically significant difference in reproductive status or menstrual cycle stage between groups (p=.912 and p=.788, respectively). Finally, participants in the CPP group reported more non-pelvic comorbidities (54.4% versus 31.8%, p<0.001) but did not take more medication for these (p=.085).

#### CPP phenotypes and self-reported diagnosis

Table 3 summarises the conditions reported by participants with PPP, grouped by symptom-based phenotypes and by their self-reported most impactful diagnosis. A single pelvic pain condition was reported by 134 of 417 participants (32.1%), with 124 (29.7%) reporting two pelvic conditions, 73 (17.6%) reporting three conditions, and 86 (20.6%) reporting four or more pelvic pain conditions. The most commonly reported symptom profile was consistent with chronic pelvic pain (56%), followed by endometriosis or adenomyosis (45%), GPPPD (44%), bladder pain syndrome (33%), and vulvodynia or vestibulodynia (16%). When asked to identify the condition they considered most impactful, 36.2% selected Endometriosis or Adenomyosis, followed by CPP (17.3%), GPPPD (11.1%), and bladder pain syndrome (10.8%).

**Table 3.** Details of self-reported pelvic pain conditions.

|  |  |  |
| --- | --- | --- |
| **Pelvic Pain Condition by Symptoms<sup>a</sup>** | Chronic Pelvic Pain | 240 (56%) |
|  | Endometriosis or Adenomyosis | 196 (45%) |
|  | Genito-Pelvic Pain/Penetration Disorder | 188 (44%) |
|  | Bladder Pain Syndrome | 141 (33%) |
|  | Vulvodynia or Vestibulodynia | 68 (16%) |
| **Self-Reported Most Impactful Pelvic Pain Condition<sup>b</sup>** | Endometriosis or Adenomyosis | 151 (36.2%) |
|  | Chronic Pelvic Pain | 72 (17.3%) |
|  | Genito-Pelvic Pain/Penetration Disorder | 46 (11.1%) |
|  | Bladder Pain Syndrome | 45 (10.8%) |
|  | Vulvodynia or Vestibulodynia | 45 (10.8%) |
|  | Pudendal Neuralgia | 19 (4.6%) |
|  | Other | 16 (3.9%) |
|  | Chronic Pelvic Girdle Pain | 15 (3.6%) |
|  | Primary Pelvic Floor Pain Syndrome | 5 (1.2%) |
|  | Pelvic Congestion Syndrome | 2 (0.5%) |

<sup>a</sup>Participants were able to choose multiple symptom profiles, 67.7% reported 2 or more symptom profiles
<sup>b</sup>Participants chose a single most impactful condition

#### Scale validation - Step 1: Item-level evaluation

Figure 1 outlines which data were used at every step of the analysis process. Complete data were available for 694 (97.6%) participants at T1, and 371 provided data at T2, though 7 were excluded from further analysis, leaving a final sample of 364 (98.1%). Item-level evaluation (see Supplemental Table 1) showed acceptable test–retest reliability for all items (ICC=.612 - .798) except item 7 “Without looking at or touching it, I can’t sense where my perineal area is” (ICC=0.333). This item also showed a ceiling effect, with more than two-thirds of respondents scoring maximally (66.9%). Item 7 was therefore excluded from further analysis.

**Figure 1.**
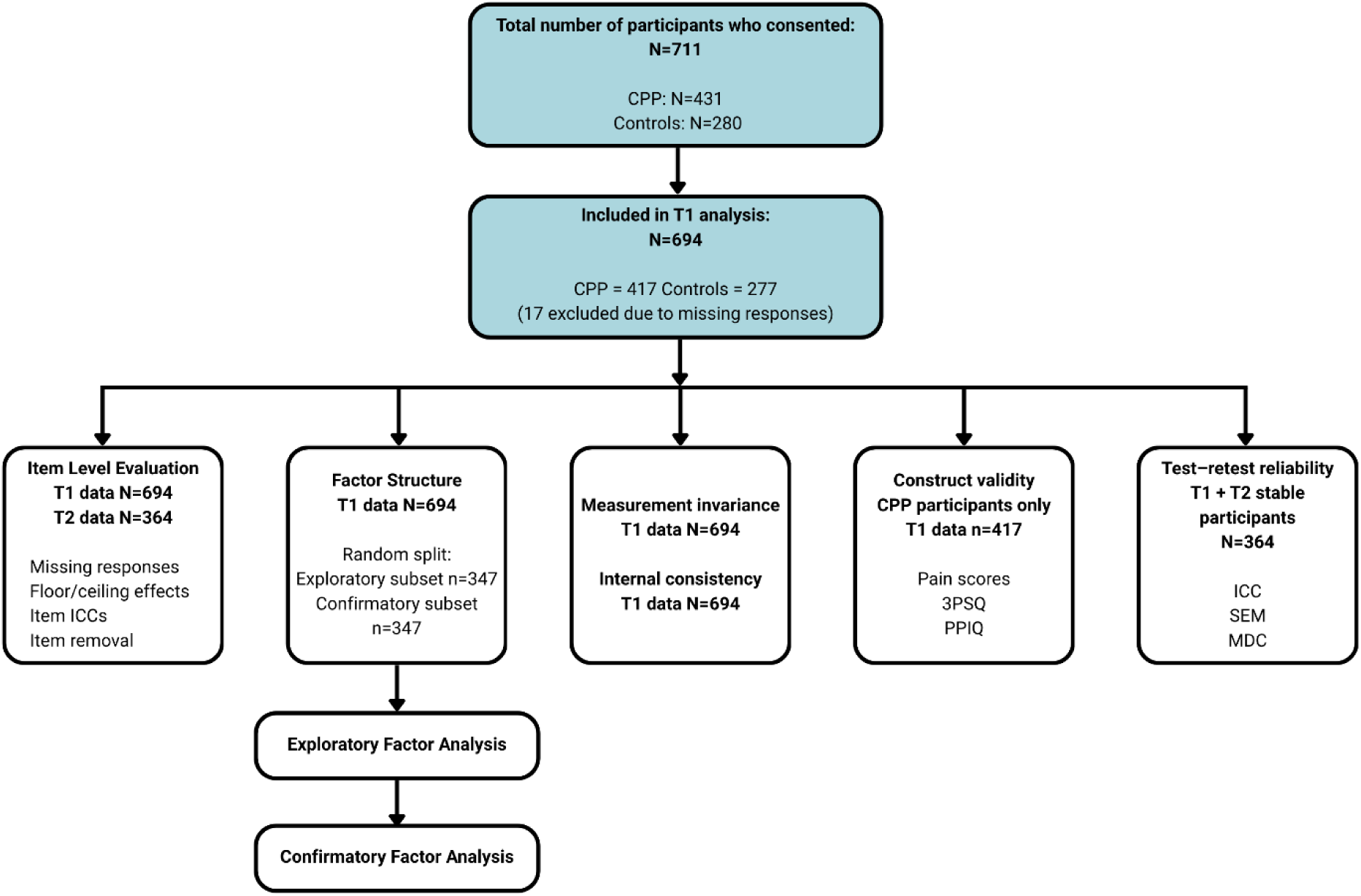
Analysis flow chart.

#### Scale validation - Step 2: Exploratory Factor analysis

Exploratory factor analysis identified two latent factors. Items one to six loaded onto one factor (range: 0.629-0.826) and items eight and nine loaded onto a second factor (range: 0.664-0.808, see Table 2 in supplementary material for more detail). Item–rest correlations (all r’s ≥ 0.30), and inter-item correlations (Factor 1: r = 0.368–0.648; Factor 2: r = 0.555) were acceptable for all items within each factor, indicating adequate homogeneity (see supplementary Tables 3 and 4). The correlation between items 2 and 3 slightly exceeded the recommended upper threshold (r = 0.719), but both were retained due to their conceptual and clinical relevance and strong prior content validity. As all item-factor loadings were sufficient (<0.3) and none of the items loaded on >1 factor, all items were retained at this stage.

#### Scale validation - Step 3: Confirmatory Factor analysis

In an initial confirmatory factory analysis run standardised item-factor loadings for all items were positive and moderate to high (.56-.85). While model fit indices showed mixed results (*χ*^2^=177.886, *p*<.001; *χ*^2^/df=4.681; CFI=.928; GFI=.938; RMSEA=.112 [.064, .163]; SRMR=0.030), inspection of modification indices revealed model fit could be improved by allowing error terms of both items 5 and 6 (MI=12.123) and items 1 and 6 (MI=11.917) to covary. On repeat analysis item-factor loadings were found to be similarly positive and moderate to high (.55 - .82), but model fit indices now improved from good to excellent (*χ*^2^=30.435, *p*=.023; *χ*^2^/df=1.790; CFI=.988; GFI=.978; RMSEA=.048 [.017, .075]; SRMR=0.0318). Figure 2 presents the resulting final overall model.

**Figure 2.**
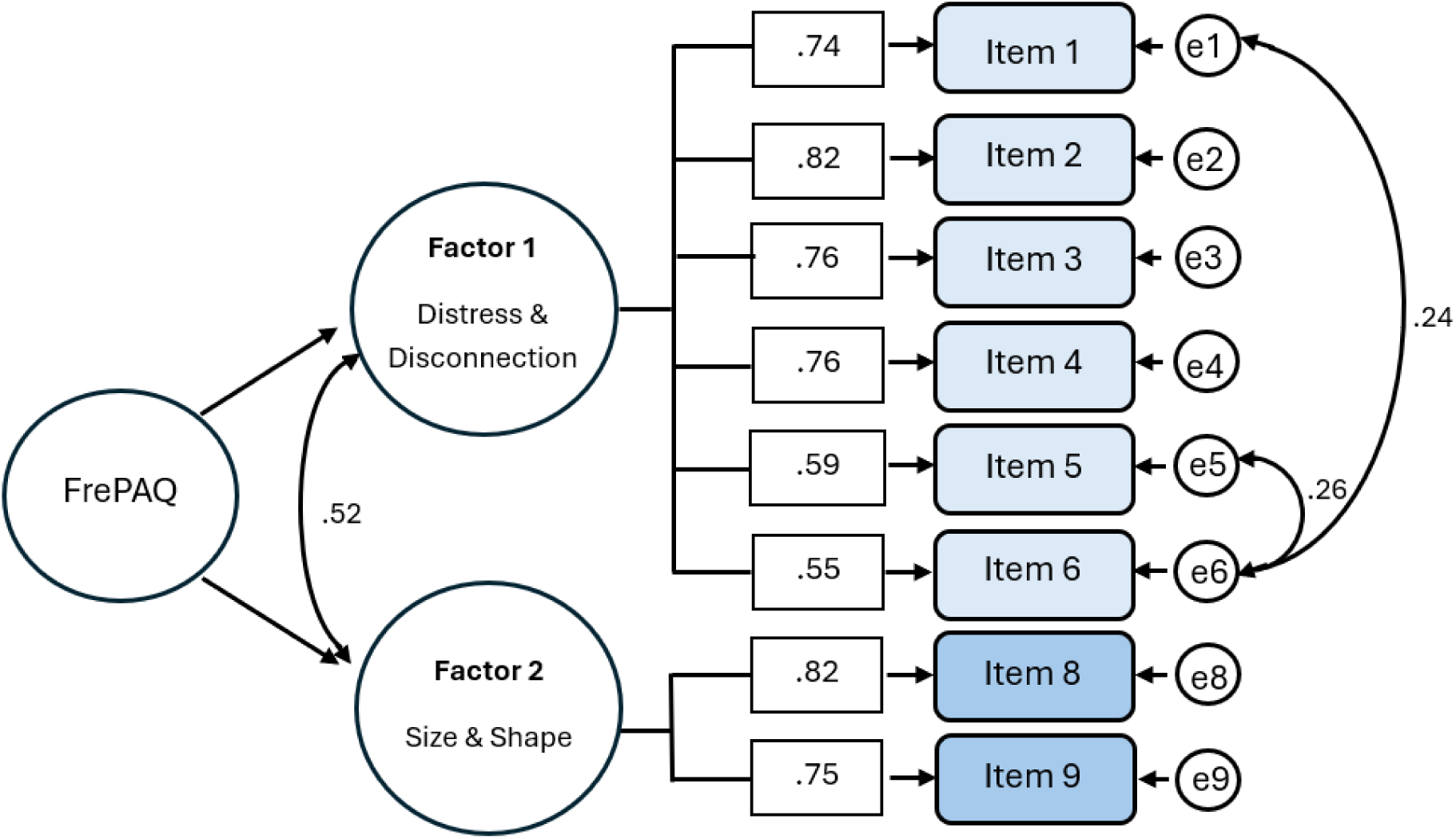
Final overall model produced by the confirmatory factor analysis. Shown are the standardised item–factor loadings, the covariances between the residual error terms for Items 1 and 6 and for Items 5 and 6, and the covariance between Factor 1 (D&D) and Factor 2 (S&S).

Discussion with the study steering group led to application of the labels “Distress & Disconnection” (D&D) for Factor 1, and “Size & Shape” (S&S) for Factor 2, and the reasons for this are further examined in discussion section.

Further tests of measurement invariance found no breach of predetermined invariance limits (CFI>.90, RMSEA and SRMR <.08, ΔCFI<-0.010, ΔRMSEA<0.015, scalar/metric invariance ΔSRMR<0.030/<0.010), indicating that the scale structure is similar across different subgroups, i.e. people with and without pelvic pain (see supplementary Table 5).

#### Scale validation - Step 4: Reliability, measurement error, and construct validity

Table 4 summarises key results. Internal consistency was good for both PPP (α=.838) and control (α=.819) groups, when applied to total scale scores of T1 data for both D&D and S&S subscales. Test–retest analysis for D&D total scores demonstrated good reliability (ICC =0.863, 95% CI: .833-.888), while results for S&S total scores (ICC =0.695, 95% CI: .635-.746) fell just below the cut-off for good reliability (0.7). The final validated version of the FrePAQ can be found in the Supplementary material.

**Table 4.**
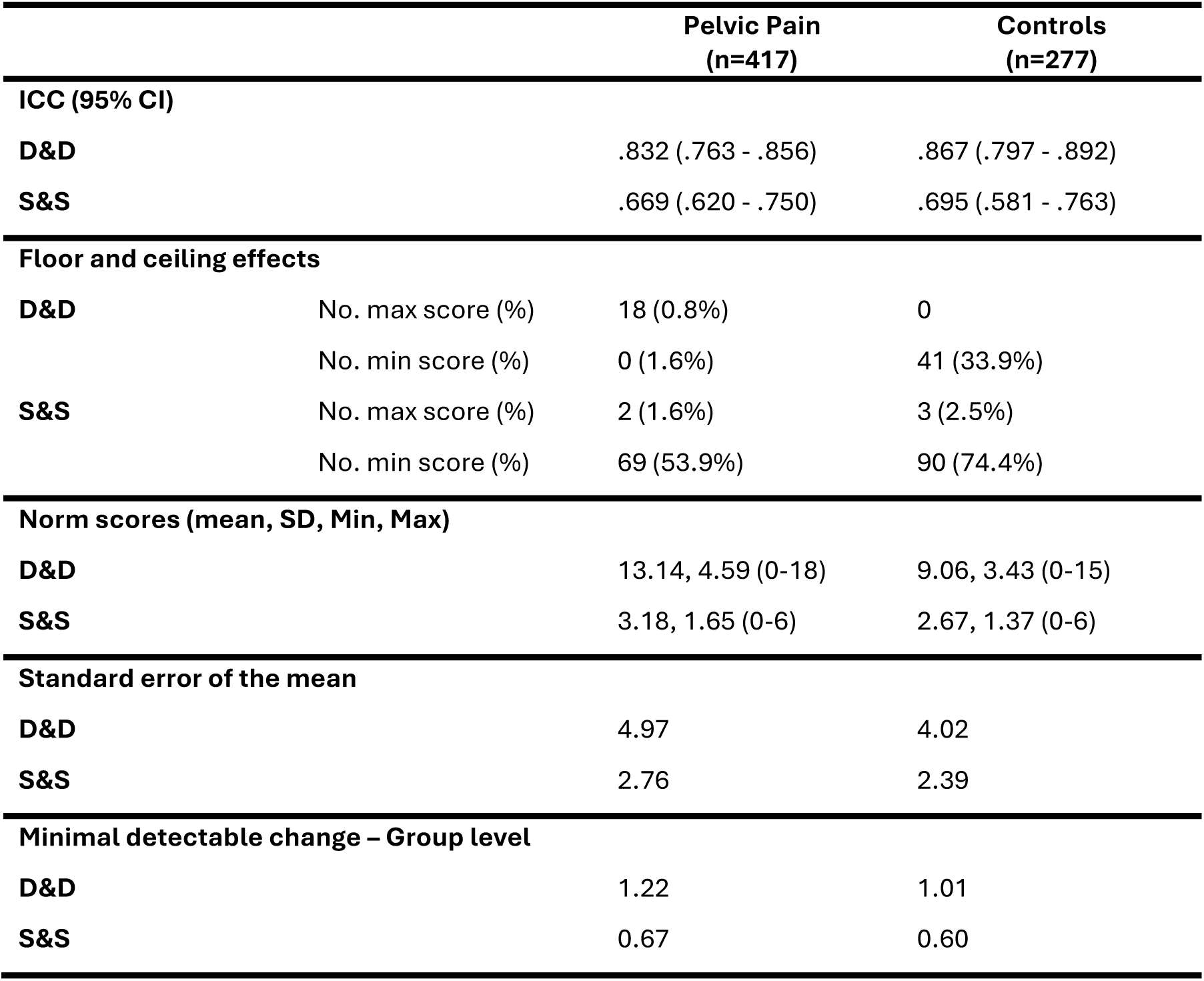
Results of measurement error, reliability and minimal detectable change.

|  |  | Pelvic Pain<br>(n=417) | Controls<br>(n=277) |
| --- | --- | --- | --- |
| **ICC (95% CI)** |  |  |  |
| **D&D** |  | .832 (.763 - .856) | .867 (.797 - .892) |
| **S&S** |  | .669 (.620 - .750) | .695 (.581 - .763) |
| **Floor and ceiling effects** |  |  |  |
| **D&D** | No. max score (%) | 18 (0.8%) | 0 |
|  | No. min score (%) | 0 (1.6%) | 41 (33.9%) |
| **S&S** | No. max score (%) | 2 (1.6%) | 3 (2.5%) |
|  | No. min score (%) | 69 (53.9%) | 90 (74.4%) |
| **Norm scores (mean, SD, Min, Max)** |  |  |  |
| **D&D** |  | 13.14, 4.59 (0-18) | 9.06, 3.43 (0-15) |
| **S&S** |  | 3.18, 1.65 (0-6) | 2.67, 1.37 (0-6) |
| **Standard error of the mean** |  |  |  |
| **D&D** |  | 4.97 | 4.02 |
| **S&S** |  | 2.76 | 2.39 |
| **Minimal detectable change – Group level** |  |  |  |
| **D&D** |  | 1.22 | 1.01 |
| **S&S** |  | 0.67 | 0.60 |

SEM values were similar across CPP and control groups. For D&D, SEM was 4.96 (CPP) and 4.02 (control), representing 20.6% and 16.7% of the score range, respectively. For S&S, SEM were 2.76 (CPP) and 2.39 (control), corresponding to 34.4% and 29.8% of the range, indicating moderate error. For CPP, MDC values for D&D were 1.22 (group level) and 13.77 (individual), and for S&S were 0.67 and 7.62, respectively.

Construct validity was examined by testing correlations between FrePAQ scores and measures of pain intensity and psychological distress using 3PSQ scores, and functional impact using PPIQ scores. As hypothesised, FrePAQ scores were positively associated with these related constructs (see supplementary Table 6 for more details). Specifically, D&D showed moderate correlations with anxiety/depression (r = .443, p < .001) and functional impact (r = .295, p < .001), and weak–moderate correlations with average pain intensity (r = .234, p < .001) and maximum pain intensity (r = .208, p < .001). However, S&S demonstrated weak associations across all comparator measures (r = .172–.226, all p < .001).

### Discussion

The aim of this study was to evaluate the psychometric properties of the FrePAQ. Overall, the findings indicate that it is a psychometrically sound measure of perineal specific BPD in women with CPP, with a stable two-factor structure, acceptable reliability, and construct validity.

In contrast to our findings, the majority of Fremantle body awareness questionnaires appear to measure one underlying construct of BPD, irrespective of body region or language. Fifteen of 23 validation studies of the Fremantle questionnaire for the back^47,54,60–66^, neck^67–69^, knee^54,70,71^, shoulder^55,72^, and for ‘general’ use^73^, have found a unidimensional underlying construct. In body regions with relatively simple functional roles in movement and daily life, where the area is easily visualised and perception relies less on interoceptive processing and has less significant sociocultural meaning, features of BPD may converge and present as a single factor. However, the perineum has a complex role in intimacy, sexuality, continence, and reproduction. It is also less visually accessible, meaning perception relies more heavily on interoception, and is shaped by strong cultural norms and expectations. In this context, the emotional and cognitive aspects of BPD may be sufficiently distinct to form a separate category. The emergence of a two-factor structure may partly reflect the nature of CPP, which commonly involves pain across multiple pelvic regions rather than a single localised body area, as targeted by other Fremantle questionnaires.

The two-factor model identified reflects theoretically meaningful dimensions of BPD, suggesting that BPD in CPP may be associated with two related but potentially distinct processes: (1) disturbances in the affective–cognitive experience of the body, and (2) alterations in perceptual representation. The first factor, ‘distress and disconnection’ (D&D), may reflect both the emotional and cognitive experience of the body, particularly altered proprioceptive awareness of pelvic floor muscle function, action, and position, that constitutes a loss of agency or disconnection with the painful region. Three items in this factor may also indicate psychological distress and functional neglect-like symptoms associated with BPD, manifesting as a sense of disconnection^74^. This finding is consistent with theoretical constructs identified in other chronic pain conditions, including neglect-like symptoms, impaired proprioceptive awareness, and modified perceptions of body ownership^8^. The associations between D&D and measures of psychological distress and pain-related functional impact suggest that this factor extends beyond merely altered sensory processing. Rather, it may capture affective–cognitive aspects of body perception, reflecting how bodily experiences arising from the pelvic region are experienced, interpreted, and emotionally appraised.

We propose that the second factor, “size and shape” (S&S), may reflect altered somatic representation experienced as distortions in perineal-region size, shape, or asymmetry. These types of perceptual disturbances have been repeatedly suggested in other chronic pain conditions such as low back pain and complex regional pain syndrome^75–81^. The potential utility of this factor is further discussed below.

Contemporary models of pain support this distinction between affective cognitive disturbances and somatic distortions. Both the Embodied Predictive Processing (EPP) model ^82^ and Fit for Purpose model^83^ conceptualise pain as arising from an ongoing interaction between bottom–up sensory input and top–down predictions about the body. Within these frameworks, perineal perception is understood not as a direct interpretation of sensory data, but as an inference shaped by prior beliefs, contextual meaning, and interoceptive signals. These models propose that body perception involves multiple interacting levels of representation, including perceptual inferences about body properties such as size, shape, and position, alongside affective and cognitive evaluations related to threat, ownership, and control. From this perspective, the two-factor structure of the FrePAQ may tentatively reflect different aspects of body perception; the S&S factor may more closely capture perceptual judgements about bodily properties, whereas the D&D factor may more broadly reflect affective and cognitive evaluations of bodily experiences arising from the pelvic region.

The IS&S factor demonstrated low overall scores, substantial floor effects, and slightly lower test–retest reliability than D&D. Nevertheless, its retention was considered justified due to its potential conceptual and clinical relevance. Although distortions in perceived size and shape appeared relatively uncommon, any non-zero S&S score may still indicate a potentially meaningful alteration in body representation, particularly when interpreted alongside D&D scores and pain and functional impact, rather than as a standalone indicator. The weaker associations with pain intensity, psychological distress, and functional impact, suggest that it may capture a more discrete aspect of BPD not fully represented by generic clinical scales. However, its lower reliability may also reflect that perceptions of size, shape, or position are more sensitive to short-term changes in bodily state or attentional focus, and may therefore reflect current perceptual experience rather than a stable trait-like construct. Overall, further research is needed to clarify its reliability, stability over time, and clinical utility across populations and contexts.

Measurement invariance confirmed that the FrePAQ measures the same latent constructs in both CPP populations and controls, suggesting that healthy controls can serve as a valid reference point to meaningfully interpret the extent and nature of BPD in pelvic pain. This supports the potential use of the measure for identifying individuals at risk, tracking changes over time, and evaluating treatment effects. More broadly, demonstrating that BPD differs between those with and without pelvic pain provides empirical support for its relevance as a clinically meaningful feature of the condition, rather than a non-specific or incidental finding.

#### Strengths and limitations

A key strength of this study is the large international sample of women with CPP alongside a substantial control group, which allowed for both a robust psychometric evaluation and meaningful comparison between clinical and non-clinical populations. Several analytic approaches following COSMIN guidance were used to provide a comprehensive assessment of the scale’s properties. Notably, this is the first Fremantle body awareness questionnaire to undergo full invariance testing, increasing confidence that the FrePAQ measures the same constructs across groups. Including a test–retest subsample further allowed assessment of score stability over time.

Several limitations should also be acknowledged. A portion of diagnoses were self-reported rather than clinically confirmed, which restricts the ability to confidently explore differences between specific pelvic pain conditions. However, diagnostic uncertainty is a recognised issue in CPP, with variability in criteria and a high proportion of individuals remaining undiagnosed. This means this sample may more accurately reflect a real-world clinical population than a strictly diagnosis-confirmed sample.

The test-retest interval was relatively short, providing evidence of short-term but not longer-term stability. Although measurement error was acceptable at the group level, the relatively higher error associated with the S&S factor, likely due to its smaller number of items and restricted score range, may limit its utility for longitudinal monitoring and should be explored in future research.

### Conclusion

This study shows that the FrePAQ is a psychometrically robust measure of altered perineal area perception in women with CPP. The two-factor structure comprises ‘distress and disconnection’ and ‘size and shape’ constructs that capture both affective–cognitive and perceptual elements of BPD. The FrePAQ demonstrated good reliability, measurement precision, and meaningful associations with clinical and psychological outcomes, indicating that it measures constructs relevant to the lived experience of pelvic pain rather than general distress or pain intensity alone. The confirmation of measurement invariance supports its use in comparing clinical and non-clinical groups. Overall, the FrePAQ represents a valuable tool for future research and may assist clinicians in identifying and addressing BPD as part of a comprehensive approach to managing CPP. In clinical practice it may be a useful tool to highlight specific features of altered body perception, inform treatment planning by distinguishing between perceptual and affective–cognitive components, and provide a means of monitoring change over time.

## Data Availability

All data produced are available online at OSF.

https://osf.io/utzjd/overview?view_only=390feb2cd84b49deaddf4341aeb508f4

## Author Statement

**Jilly Bond:** Conceptualization, Methodology, Formal analysis, Validation, Investigation, Data curation, Visualization, Writing – original draft, Writing – review & editing, Project administration, **Neil E O’Connell:** Investigation, Writing – review & editing, Supervision, **Benedict M Wand:** Methodology, Writing – review & editing, **K. Jane Chalmers:** Writing – review & editing, **Elmar Kal:** Methodology, Validation, Formal analysis, Data curation, Writing – review & editing, Supervision.

## Acknowledgements

The research team would like to thank the informal research advisory team for their support and guidance in the development of this study: The Vulval Pain Society, Bladder Health UK, The MASIC Foundation, Sheren Gaulbert, Dr Adanna Okeahialam, Megan Rae and Susannah Fraser.

## Disclosures

JB is a co-director of the Pelvic Pain Network and lectures regarding physiotherapy for pelvic pain.

NOC is a member of the Cochrane Central Editorial Board. Between 2020 and 2023 Neil was Co-ordinating Editor of the Cochrane Pain, Palliative and Supportive Care group, whose activities were funded by an infrastructure grant from the UK National Institute of Health and Care Research (NIHR). He is the Chair of the International Association for the Study of Pain (IASP) Methodology, Evidence Synthesis and Implementation special interest group (MESISIG) and has held a grant from the ERA-NET Neuron Co-Fund.

BMW, KJC and EK have nothing to declare

This study received no funding.

No AI tools were used in this study.

The protocol was pre-registered through the Open Science Framework: https://shorturl.at/M60aL

## Data Statement

Data from this study are available open access here: https://tinyurl.com/5898x5pz

## Supplemental Material

### Study Recruitment partners

The following charities and organisations disseminated the study recruitment poster.

### Charities

The Vulval Pain Society UK

Bladder Health UK

Fair Treatment for Welsh Women (FTWW)

Women Connect First

### Organisations

The Pelvic, Obstetric and Gynaecological Physiotherapy (POGP) professional network affiliated to the Chartered Society of Physiotherapy (including Northern Ireland)

International Organisation of Physiotherapists in Pelvic and Women’s Health (IOPPWH)

Irish Society of Chartered Physiotherapists Pelvic Health and Continence group

The American Physical Therapy Association Academy for Pelvic Health

Canadian Association of Physiotherapy Division of Pelvic Health

Endometriosis UK Endometriosis South of Scotland Endometriosis Australia

The Endometriosis Network Canada

Pelvic Pain Support Network UK

### Background questions given to all participants

Age, country of residence, whether they were born in country they reside, self-defined ethnicity, self-defined sexual orientation, highest level of schooling completed, parity, menopause status, current point of menstrual cycle, presence of non-pelvic medical conditions, list of non-pelvic medical conditions, number of medications taken for non-pelvic medical conditions, partaking in regular physical activity, number of hours of exercise per week, presence of pelvic pain,

### Further background questions given only to participants reporting pelvic pain

Average pain in last 24 hours, maximum pain in the last 24 hours, main or most impactful condition, presence of other pelvic pain conditions, whether a formal diagnosis has been given, symptom profiles for pelvic pain conditions, number of years of symptoms, number of years since diagnosis, number of medical professionals seen regarding pelvic pain condition, number of surgeries for pelvic pain condition, analgesic medications taken

### The 9-item Fremantle Perineal Awareness Questionnaire

The pelvic area is the whole lower part of the trunk, including the lower back, pelvis, hips, and bottom. The **perineal area** is a small part of this located **between the legs**. It covers the area from the pubic bone to the tailbone and between the two sit bones. This area includes the genitals, pelvic floor muscles, anal area, and skin (perineum). Functions of this area include bladder and bowel control (peeing and pooing), sexual function, supporting pelvic organs, and assisting movements of the body (sitting, walking, running, and jumping). People with problems in this area often feel symptoms in other areas of the pelvis, however, this questionnaire is specifically focusing on the perineal area and how that area feels to you.

Problems in this area are common but may be difficult to discuss with people in your life, including your healthcare providers. One issue is that the way your perineal area feels can change. This questionnaire assesses how changes in your perineal area can impact your function or cause you distress.

Using the following scale, answer each question to let us know how this area feels to you:

0 = **Never** feels like that

1= **Occasionally** feels like that

2= **Often** feels like that

3= **Always** feels like that

|  | Never | Occasionally | Often | Always |
| --- | --- | --- | --- | --- |
| 1. I feel disconnected from my perineal area. | 0 | 1 | 2 | 3 |
| 2. I feel distressed when I look at or think about my perineal area. | 0 | 1 | 2 | 3 |
| 3. I feel distressed when I touch, am touched, or think about being touched in my perineal area. | 0 | 1 | 2 | 3 |
| 4. Using my perineal area takes more conscious effort than I think it should. | 0 | 1 | 2 | 3 |
| 5. I feel as if the muscles in my perineal area tighten or relax without my control. | 0 | 1 | 2 | 3 |
| 6. When using my perineal area, I am unaware or unsure of what the muscles in the area are doing. | 0 | 1 | 2 | 3 |
| 7. Without looking at or touching it, I can't sense where my perineal area is. | 0 | 1 | 2 | 3 |
| 8. Without looking at or touching it, my perineal area feels like it has changed in shape or size. | 0 | 1 | 2 | 3 |
| 9. Without looking at or touching it, my perineal area feels lopsided asymmetrical, or uneven. | 0 | 1 | 2 | 3 |

Total score= \_\_\_\_/27

**Table 1.** Summary of item-level evaluation of the FrePAQ.

| Item | n / %<br>missing | % min/max<br>score | ICC (95% CI) | Included? |
| --- | --- | --- | --- | --- |
| 1. I feel disconnected from my perineal area | 3 / 0.4% | 5.3 / 48.9 | .660 (.596, .716) | Yes |
| 2. I feel distressed when I look at or think about my perineal area | 6 / 0.8% | 7.9 / 49.9 | .742 (.690, .787) | Yes |
| 3. I feel distressed when I touch, am touched, or think about being touched in my perineal area | 5 / 0.7% | 8.0 / 46.6 | .772 (.725, .812) | Yes |
| 4. Using my perineal area takes more conscious thought than I think it should | 4 / 0.6% | 11.4 / 43.0 | .685 (.624, .737) | Yes |
| 5. I feel as if the muscles in my perineal area tighten without my control | 4 / 0.6% | 13.6 / 38.4 | .798 (.756, .834) | Yes |
| 6. When using my perineal area, I am unaware or unsure of what the muscles in the area are doing | 6 / 0.8% | 7.9 / 46.0 | .626 (.557, .687) | Yes |
| 7. Without looking at or touching it, I can't sense where my perineal area is | 7 / 1% | 10.8 / 66.9 | **.333 (.235, .425)** | No |
| 8. Without looking at or touching it, my perineal area feels like it has changed in shape or size | 7 / 1% | 4.5 / 67.4 | .612 (.541, .675) | Yes |
| 9. Without looking at or touching it, my perineal area feels lopsided, asymmetrical or uneven | 8 / 1.1% | 5.9 / 71.9 | .650 (.583, .707) | Yes |

NB: Item 7 highlighted in red was removed from analysis due to low reliability and a ceiling effect

**Table 2.** FrePAQ exploratory factor analysis – item-factor loadings.

| Item | Factor |  |
| --- | --- | --- |
|  | 1 | 2 |
| 1. I feel disconnected from my perineal area. | .723 | -.045 |
| 2. I feel distressed when I look at or think about my perineal area. | .670 | .168 |
| 3. I feel distressed when I touch, am touched, or think about being touched in my perineal area. | .726 | .089 |
| 4. Using my perineal area takes more conscious effort than I think it should. | .826 | -.003 |
| 5. I feel as if the muscles in my perineal area tighten or relax without my control. | .639 | -.060 |
| 6. When using my perineal area, I am unaware or unsure of what the muscles in the area are doing. | .629 | -.008 |
| 8. Without looking at or touching it, my perineal area feels like it has changed in shape or size. | -.046 | .808 |
| 9. Without looking at or touching it, my perineal area feels lopsided asymmetrical, or uneven. | .081 | .664 |

Extraction method: Principal axis factoring with oblimin rotation. KMO = .851; Bartlett's test of sphericity $\chi^2(28) = 1152.35$ , $p < .001$ . The two-factor solution explained 53.1% of total variance.

**Table 3.** FrePAQ Inter-item correlation matrix for the first factor.

|  | Item 1 | Item 2 | Item 3 | Item 4 | Item 5 | Item 6 |
| --- | --- | --- | --- | --- | --- | --- |
| Item 1 | 1.000 | .578 | .531 | .570 | .398 | .449 |
| Item 2 |  | 1.000 | .719 | .604 | .368 | .398 |
| Item 3 |  |  | 1.000 | .648 | .414 | .408 |
| Item 4 |  |  |  | 1.000 | .526 | .506 |
| Item 5 |  |  |  |  | 1.000 | .552 |
| Item 6 |  |  |  |  |  | 1.000 |

**Table 4.**
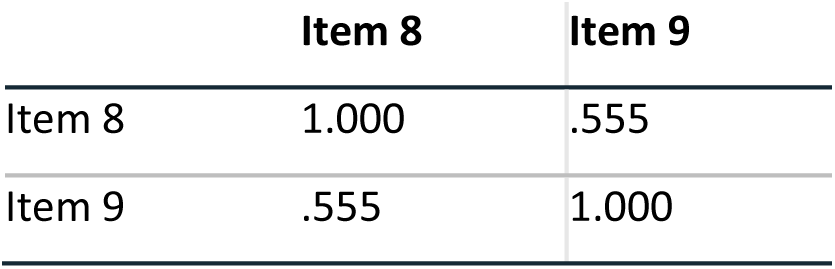
FrePAQ inter-item correlation matrix for the second factor.

**Table 5.**
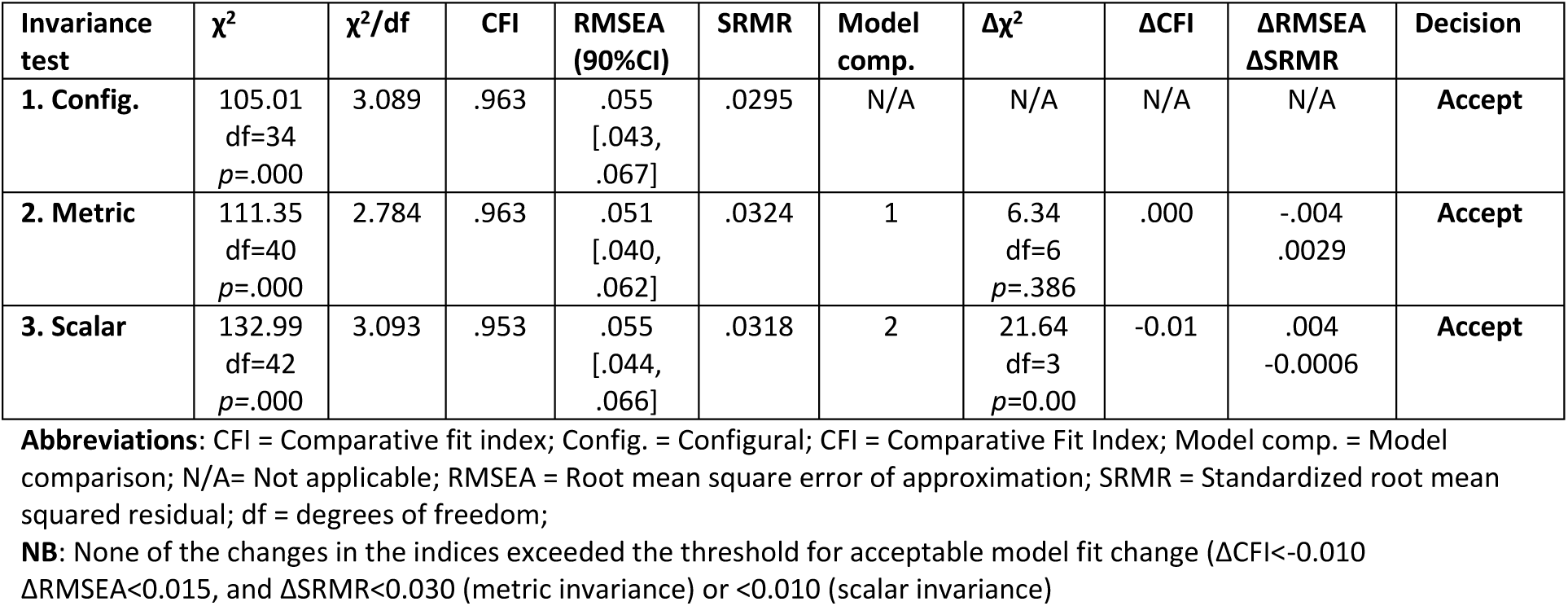
Results of measurement invariance testing.

**Table 6.**
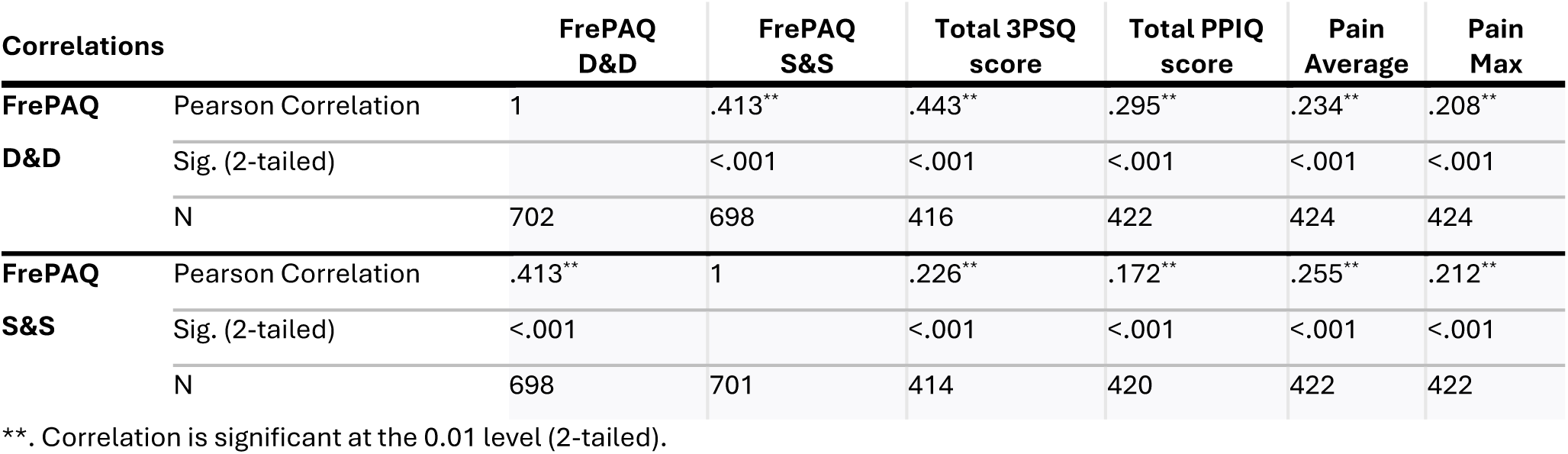
Results of construct validity testing.

### The Fremantle Perineal Assessment Questionnaire (FrePAQ)

The pelvic area is the whole lower part of the trunk, including the lower back, pelvis, hips, and bottom. The **perineal area** is a small part of this located **between the legs**. It covers the area from the pubic bone to the tailbone and between the two sit bones. This area includes the genitals, pelvic floor muscles, anal area, and skin (perineum). Functions of this area include bladder and bowel control (peeing and pooing), sexual function, supporting pelvic organs, and assisting movements of the body (sitting, walking, running, and jumping). People with problems in this area often feel symptoms in other areas of the pelvis, however, this questionnaire is specifically focusing on the perineal area and how that area feels to you.

Problems in this area are common but may be difficult to discuss with people in your life, including your healthcare providers. One issue is that the way your perineal area feels can change. This questionnaire assesses how changes in your perineal area can impact your function or cause you distress.

Using the following scale, answer each question to let us know how this area feels to you:

0 = **Never** feels like that

1 = **Occasionally** feels like that

2 = **Often** feels like that

3 = **Always** feels like that

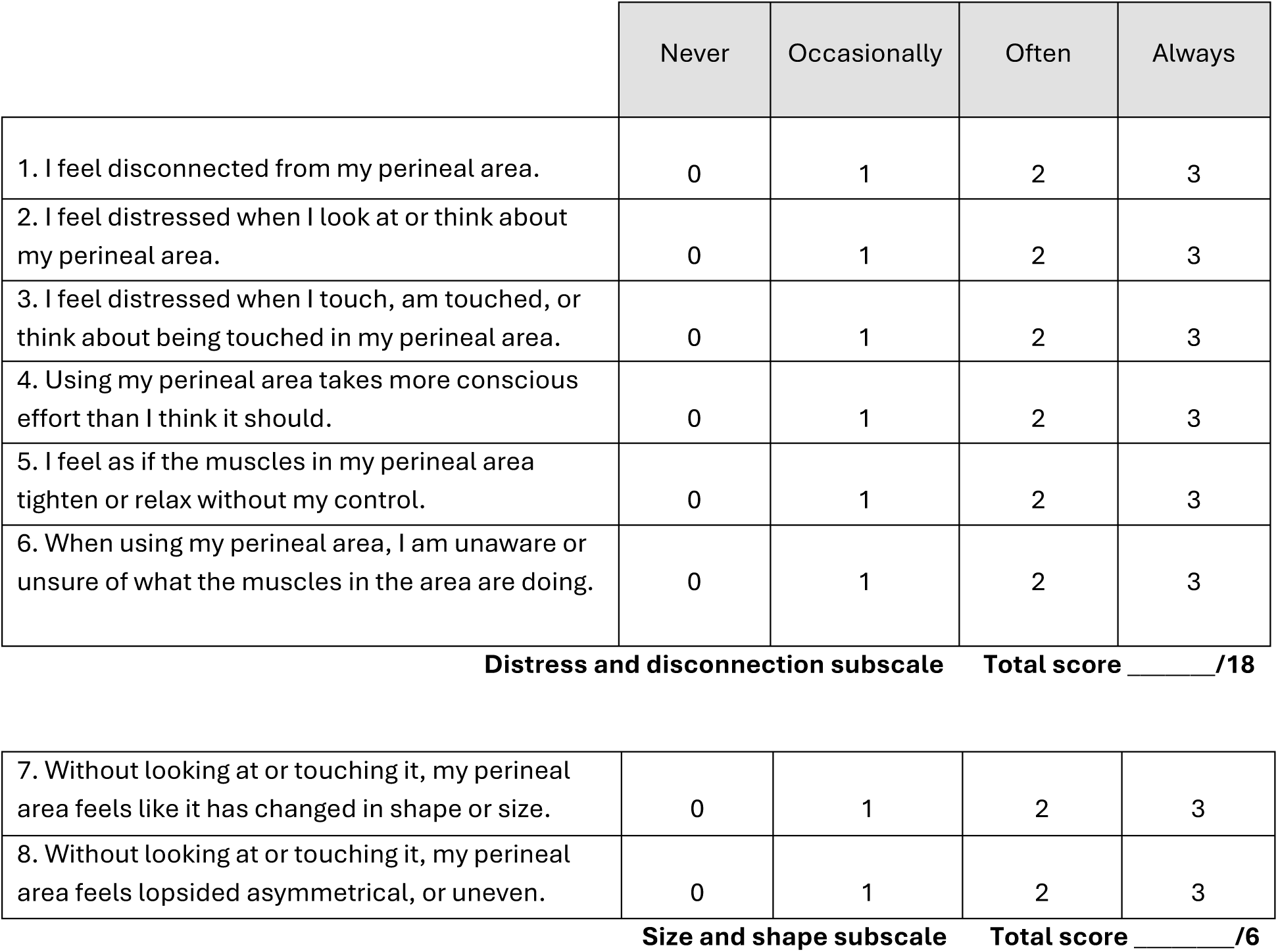

|  | Never | Occasionally | Often | Always |
| --- | --- | --- | --- | --- |
| 1. I feel disconnected from my perineal area. | 0 | 1 | 2 | 3 |
| 2. I feel distressed when I look at or think about my perineal area. | 0 | 1 | 2 | 3 |
| 3. I feel distressed when I touch, am touched, or think about being touched in my perineal area. | 0 | 1 | 2 | 3 |
| 4. Using my perineal area takes more conscious effort than I think it should. | 0 | 1 | 2 | 3 |
| 5. I feel as if the muscles in my perineal area tighten or relax without my control. | 0 | 1 | 2 | 3 |
| 6. When using my perineal area, I am unaware or unsure of what the muscles in the area are doing. | 0 | 1 | 2 | 3 |
| **Distress and disconnection subscale** Total score \_\_\_\_/18 |  |  |  |  |
| 7. Without looking at or touching it, my perineal area feels like it has changed in shape or size. | 0 | 1 | 2 | 3 |
| 8. Without looking at or touching it, my perineal area feels lopsided asymmetrical, or uneven. | 0 | 1 | 2 | 3 |

**Size and shape subscale** Total score \_\_\_\_/6

